# Sanitation-Hygiene Practices and Social Norms in Community-led Total Sanitation for Sustainability of Open Defecation Free Status: A Survey of Suna West Sub-County, Migori County, Kenya

**DOI:** 10.1101/2025.08.01.25332764

**Authors:** Naomi R. Aluoch, Collins O. Asweto, Patrick O. Onyango

## Abstract

**Background:** Community-led total sanitation (CLTS) has been used to stir sanitation-related behaviour change and attain open defecation free (ODF) status. CLTS interventions suffer high rates of reversion such that their gains are unsustainable in most contexts.

**Objective:** This study aimed to determine the role of sanitation hygiene practices and social norms on open defecation free status in Suna West Sub County.

**Methodology:** A cross-section study design was employed using questionnaire and observation checklist to collect data from 384 households.

**Results:** Results revealed that 66.1% households had partially reverted to non-ODF status. The sanitation-hygiene practices associated with being ODF includes: use of elevated racks (AOR=0.81; CI=0.34-1.90; *p*=0.625), use of treating water (AOR=2.81; CI=(0.97-8.06); *p*=0.055), regularly clean latrines (AOR=2.96; CI=0.63-13.89; *p*=0.17), pouring of ash over the pit of the latrine (AOR=4.08; CI=1.73-9.62; *p*<0.001) and use of dug out pits for waste disposal (AOR=2.41; CI=1.02-5.68; *p*<0.045). On social norms, the study found that laws/penalties (AOR=4.15; CI=2.20-7.80; *p*<0.001) and rewards/incentives (AOR=0.17; CI=0.096-0.306; *p*<0.001) had less odds of being ODF. Moreover, odds of being ODF was low for households who reported that construction/maintenance materials were expensive (AOR=0.17; CI=0.84-2.79; *p*=0.169), that it was embarrassing to people defecate in the open (AOR=0.060; CI0.19-1.23=; p<0.129) and that it is okay to defecate in bushes/rivers/dams (AOR=0.623; CI=;0.329-1.179 p<0.146).

**Conclusion:** The results of this study show partial reversion to non-ODF status in previously certified villages. However, households that sustained ODF status had several sanitation hygiene practices. Interestingly, households that displayed social norms were less likely to be ODF. Overall, the findings of the present study demonstrate that the CLTS process failed to instil social norms around proper sanitation to inspire community collective action thus little influence on sustainable behaviour change. The findings of this study therefore highlight the need to enhance good hygiene sanitation practices, while instilling social norms to inspire community collective action.

## INTRODUCTION

According to the World Health Organisation (WHO), roughly 842,000 lives are lost in low- and middle-income countries annually as a consequence of inadequate water, hygiene and sanitation (1). Indeed, poor sanitation has been connected to infections such as (2) diarrhoeal diseases, nematode infections and environmental enteropathy (EE). In Kenya, for instance, diarrhoea claims the lives of roughly 3,100 children annually and trachoma, schistosomiasis are health problems linked to poor sanitation. In part, the burden of these diseases is attributed to open defecation that exposes a large part of the population to sanitation-related diseases (3).

It is in light of such negative impacts of poor sanitation that the Government of Kenya adopted Community-led Total Sanitation (CLTS) as a strategy to improve sanitation. Community-led Total Sanitation was introduced by Plan International Kenya in 2007 and was approved by the then Ministry of Public Health and Sanitation (MOPHS) as a key framework for promoting hygiene and sanitation at the household level. In 2011, MOPHS established CLTS as the national strategy for ensuring rural sanitation and set a national target to reduce open defecation (4).

Community led total sanitation involves three main steps: pre-triggering, triggering and post-triggering(5). At the pre-triggering stage the community members are mobilised for triggering process in which community members are able to see the reality of mass defecation and how it affects the community negatively (6). Finally, the post-triggering phase involves follow-up aimed at verification and certification of the villages as open defecation free(7).

There has been a lot of research on why people do what they do, what influences their actions. In the recent sanitation interventions and community-led total sanitation, key emphasis has been on establishing social norm around unacceptability of open defecation (6). The process of CLTS has been known to cause peer pressure by invoking emotions such as shame and disgust and change perceived social norms to establish open defecation as being socially unacceptable(8). Social norms and how strong they are, has been found to greatly influence sustainable behaviour change in relation to sanitation practices. Where positive social norms were well rooted, the chances of achieving sustainable behaviour change were higher(9).

It is important to understand why people engage in open defecation for failure to do so has led to failed interventions(10). Open defecation has been known to be an independent action, that is, people engage in open defecation because they believe it meets their needs and that it is not harmful to them and others, it is a custom rather than a social norm (11). However, to eliminate open defecation in a particular group, there is need to create a social norm promoting latrine use and maintenance. People will need to think that their reference network think they should use and maintain latrines and them believing in that will motivate them to engage in open defecation-free behaviour(10).

The results of a study on the sustainability of ODF status in Kenya conducted by (12) revealed that the sustainability of ODF achievements remained a major concern with over 70% of villages that had received partial or full ODF status reverting to non-ODF status. Among the factors that demotivate community members from using a latrine after becoming ODF relates to physical aspects of the latrine (such as lack of privacy and fear of the latrine collapsing) and sharing a latrine with other people (14). In addition, slippage from ODF status has also been linked to collapse or poor structural integrity of latrines as well as unsustainable behaviour change following sanitation-related interventions (14). The objective of this study was to investigate the ODF status of households in ODF certified villages of Suna West Sub County and to determine association between ODF status with sanitation-hygiene practices and social norms in households of Suna West Sub-County, western Kenya.

## MATERIALS AND METHODS

### Study Area

The study was done in Suna West Sub-County in Migori County which has a population of 117,539 with a density of 406 persons per km^2^ and 29,251 households. The sub-county, one among eight others, has four wards; Ragana-Oruba, Wasweta II, Wiga and Wasimbete wards. It is bordered by Kuria West sub-county to the south-east, Nyatike sub-county to the west, Suna East Sub-County to the north-eastern side and Tanzania to the south-west. Migori county has poverty level measured at 32.0 (2016) and is listed among the counties with the most income inequality as measured by the gini coefficient. Migori county’s a gini index is 0.464(15).

### Study Design

A cross-sectional study design was used across two wards, Ragana-Oruba and Wasweta II, that were purposively chosen for having attained ODF status at least one year to the study which was carried out from 17^th^ to 21^st^ July, 2020.

### Target population

The unit of analysis was the household with the targeted participants being the household heads. The sample size for households featured in the study was determined using the statistical formula as used by Fisher .(16). The households that had participated in the initial verification were included and any child-headed household was excluded. The data collection was done across 384 households identified through simple random sampling of the households as found in MOH 513 which is the household register maintained by the Community Health Volunteers (CHVs).

### Sampling Design

The sample size for households in the village featured in the study was determined using Fisher’s formula; the standard deviation set at 1.96, which corresponds to 95% CI. The proportion of desired population was assumed at 0.50, the margin of error 0.05. We used the initial sample size of 384 to capture the true variability of the population.

### Data Collection Tools and Procedure

The survey tools were developed in line with the constructs of the key questions that we sought to answer with reference to previously validated instruments that were reviewed by my supervisors and peers. Once that was done, we pre-tested the tool and adjusted any questions that we sought to answer. Cronbach’s alpha was computed to test for reliability of data collection instrument and output yielded result of an internal consistency 0.7.

Validated structured questionnaire and observation checklist were used in this study. Observation checklist was used to collect information to corporate or refute claims made by respondents in the questionnaires.

The researcher provided scientific oversight of this study including training and technical support for the research team and oversaw the development of coding frame and data analysis. Under supervision, the research assistants (RAs) helped in the recruitment and obtaining informed consent from the participants to take in the part in the study.

The tools were provided in English.

### Study Variables

#### Dependent Variables

Dependent variables were measured as follows: a) access to a latrine - availability of Individual latrine, shared latrine/neighbours; b) privacy - availability of door or some form of barricade provided for each superstructure; c) Squat hole cover – a barrier to prevent movement of flies in and out of the latrine hole d) hand washing facility - availability of tap/leaky tin near latrine with water inside, soap/ash available; e) no exposed faeces – via simple transect walk, no visible faeces should be found within the surrounding of the home. For a household to be ODF, all the 5 parameters must be met and the absence of any one would be an indication of partial reversion to non-ODF status while absence of all the five indicators would reflect full reversion to non-ODF status of the household.

#### Independent Variables

For independent variables, the frequency of variables such as treating water (boiling or use of chemicals), covering food using lid over cooking pots when cooking and during storage, using elevated racks to hold utensils off the ground while drying, regular cleaning of latrine, application of ash around & the squat hole of latrine, and using dugout pit for waste disposal were measured using Likert scale: always, most of the time, sometimes, rarely, not at all. Scores were aggregated into two - Yes (always, most of the time, sometimes) and no (rarely, not at all).

While variables such as Care of the family - Empirical & normative expectation regarding health of the family, Shame/disgust/fear/pride Regrettable occurrence/ unpleasant emotion that cause a feeling of resolution, Cultural/social/religious beliefs - Person’s belief alignment as pertaining culture, society and religion, Laws/penalties - Rules within a given set up and punishment imposed for breaking the set rules, Need to improve things in the family - Empirical & normative expression of obligation to make things better, Follow ups and support - The subsequent actions following CLTS and material assistance for the same, Rewards/incentives - Some form of payment given in recognition of work done or to stimulate greater output, and Peer pressure - The empirical & normative expectation regarding consistent latrine use were measured using Likert scale: strongly disagree, disagree, neutral, agree and strongly agree. The scores were aggregated into two; Yes (agree and strongly agree), No (strongly disagree, disagree, neutral).

#### Statistical Analysis

Summation for the observed non-negotiable indicators was done to determine the ODF status. Wilcoxon signed-rank test was used to determine if there was a significant difference in ODF status as at the time of the study and verification. Chi-square test of independent was used to determine association between sanitation and hygiene practices, social norms and ODF status and binary logistic regression was done to determine the relationship between sanitation hygiene practices, social norms and ODF status.

While the outcome variable was highly prevalent, raising concerns about the potential for odds ratios to over-estimate the true association compared to risk ratios, we chose to use logistic regression as it offers a robust framework for adjusting for multiple confounders and provides a straightforward method for hypothesis testing. Nevertheless, we interpreted the results with caution, to effectively utilize established statistical methodologies and maintain consistency with previous research in similar contexts.

#### Ethical considerations

Ethical approval for the study was granted by the Maseno University Ethics Review Committee; Ref: MSU/DRPI/MUERC/00821/99 and National Commission for Science Technology and innovation, Kenya. To protect the privacy and anonymity of participants, their personal identifying information was not recorded in the questionnaires or checklists and households were reported as numbers. The data was coded to secure the privacy and anonymity of participants. Further, there was disclosure regarding the nature of the study, its purpose, what it involves, the procedures to be done, risks-if any by taking part in the study and since the study did not involve experimental drugs or procedures used in the study, the study posed no significant risk to participants.

#### Informed consent

Written informed consent was gotten from the participants. They were also informed that taking part in the study was out of free will and that they were free to withdraw from the study at any time.

## RESULTS

### Socio-demographic characteristics of study participants

Of the 384 participants, 62.8% were females, 73.4% were aged 25-59 years, while 58.6% and 31.8% had primary education and secondary education respectively. On socio-economic status, three-quarter (75.3%) of households had $0-38.9 monthly income. A half (53.1%) of the households had 0-5 years old child and 27.1% had at least one member with a disability or chronic illness. Socio-demographic characteristics of the study participants are summarized in Table 1.

**Table 1.**
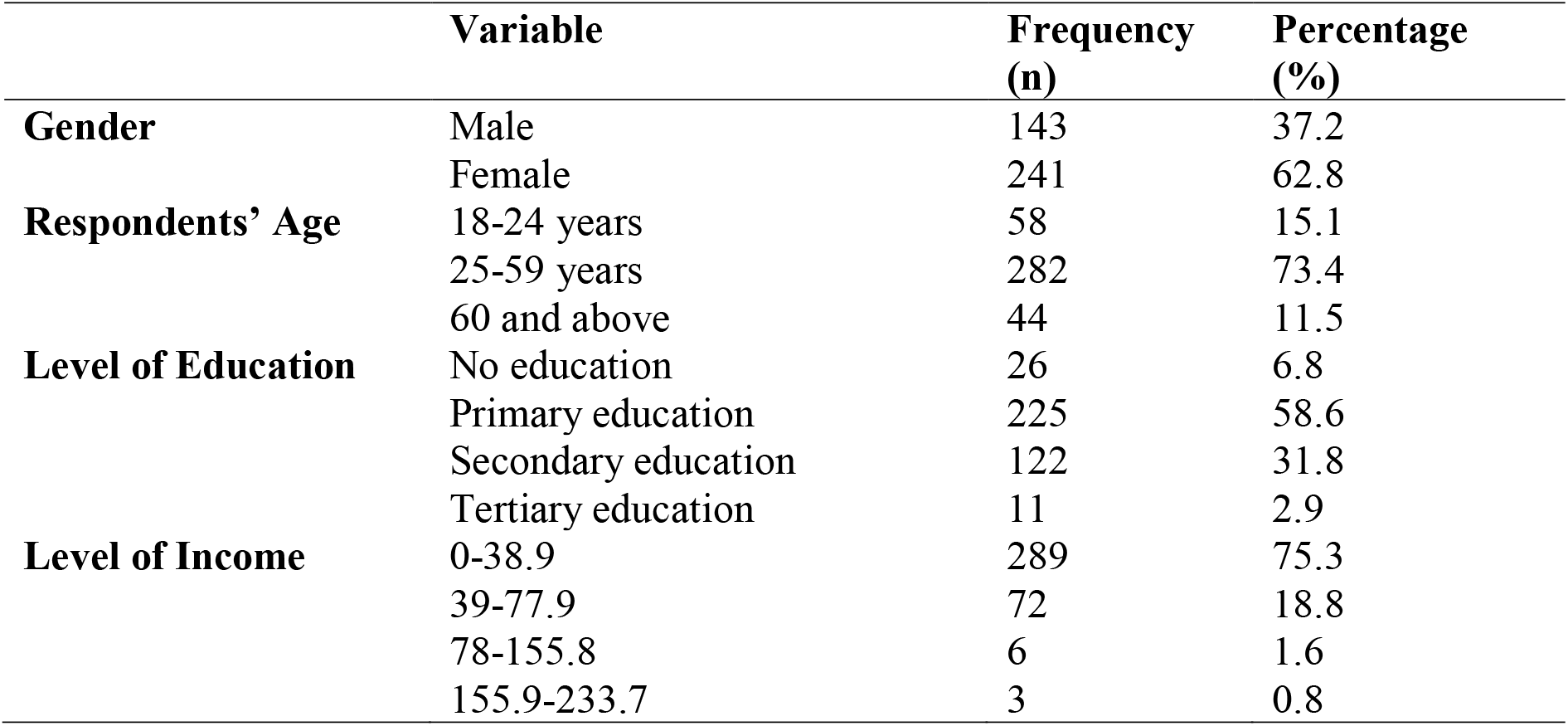
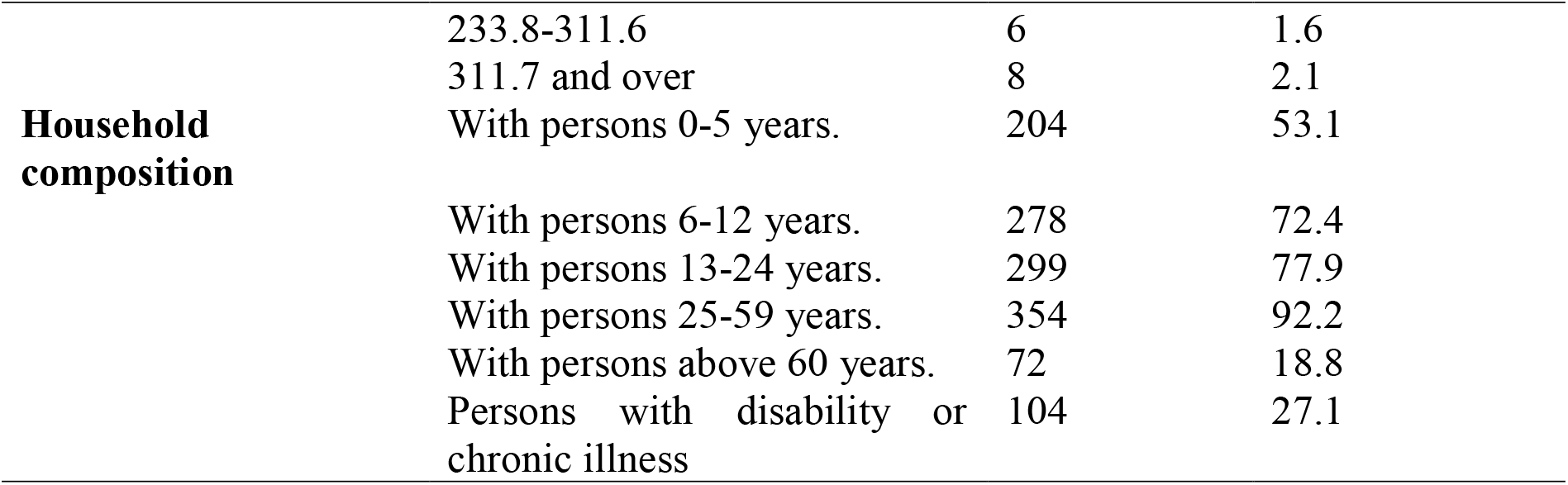
Socio-demographic characteristics of study participants from Suna West Sub-County, Kenya. Table legend 1: The average exchange rate as at the time of the study was **KES 128.37 for a dollar**

### Open Defecation Free Status

On ODF status, only 33.9 % (n=130) were found to be ODF one year after certification. When the indicators were analysed singly, it was observed that access to latrine and no exposed faeces were at 100%; with 95.3% (n=366) owning individual latrines while the remaining 4.7% (n=18) reporting to use shared latrines, Table 2.

**Table 2.** Results on ODF indicators.

| **Indicator** | **Median percentage (%)** | **P value** | **No. of villages reporting 100%** |
| --- | --- | --- | --- |
| Access to latrine | 100 | 1.0 | 13 (100%) |
| Squat hole cover present | 63 | 0.002 | 0 (0%) |
| Privacy | 82.4 | 0.002 | 0 (0%) |
| Hand washing facility | 82.4 | 0.004 | 2 (15%) |
| No exposed faeces | 100 | 0.056 | 8 (61.5%) |

### Association between sanitation and hygiene practices and open defecation free status

This analysis determined the association between various household sanitation practices and the likelihood of being Open Defecation Free (ODF). The results show that household that pour ash into latrines had higher odds of ODF status compared to households that do not (AOR=4.082; 95% CI:1.730 – 9.615; p value = 0.001), while households that dug out pits for waste disposal had higher odds of ODF status compared to households that do not (AOR=2.410; 95% CI:1.018 – 5.682; p value = 0.045). Other practices such as treating water, using elevated racks, and regularly cleaning of latrines were not significantly associated with ODF as shown in Table 3.

**Table 3.** Association between sanitation and hygiene practices and open defecation free status in Suna West Sub-County, Kenya.

| Characteristic | ODF n (%) | OR (95%CI) | P value | AOR (95%CI) | P Value |
| --- | --- | --- | --- | --- | --- |
| **Treating water** |  |  |  |  |  |
| Yes | 123 (35.3) | 3.226(0.912–8.547) | **0.019** | 2.809 (0.978-8.065) | **0.055** |
| No | 5 (14.7) | Ref |  | Ref |  |
| **Using elevated racks** |  |  |  |  |  |
| Yes | 117 (35.7) | 2.203 (1.106-4.444) | **0.027** | 0.808 (0.343-1.901) | **0.625** |
| No | 11 (20.4) | Ref |  | Ref |  |
| **Regular cleaning of the latrine** |  |  |  |  |  |
| Yes | 128 (35.2) | 4.878 (1.122-13.513) | **0.035** | 2.959 (0.629-13.889) | **0.170** |
| No | 2 (10.0) | Ref |  | Ref |  |
| **Pouring of ash** |  |  |  |  |  |
| Yes | 121 (38.5) | 4.348 (2.083-9.091) | **<0.001** | 4.082 (1.730-9.615) | **0.001** |
| No | 9 (12.9) | Ref |  | Ref |  |
| **Dug out pit for waste disposal** |  |  |  |  |  |
| Yes | 122 (38.4) | 4.504 (2.083-9.804) | **<0.001** | 2.410 (1.018-5.682) | **0.045** |
| No | 8 (12.1) | Ref |  | Ref |  |

### Association between social norms and open defecation free status

This study found association in a number of the social norms and ODF status. For instance, individuals who were not subjected to laws and penalties were considerably more inclined to be ODF compared to those who were (AOR = 4.145; 95% CI: 2.203–7.802; p < 0.001), indicating that the implementation and enforcement of legal frameworks are not essential for encouraging ODF behaviours. Furthermore, the lack of follow-ups and support was correlated with increased odds of open defecation (OR = 1.985; p = 0.025), although the adjusted effect was not statistically significant (AOR = 2.111; p = 0.118). While follow-up support appears to have some effect, its impact weakens when accounting for other variables. Interestingly, those who did not anticipate receiving incentives were notably more likely to retain ODF status (AOR = 0.172; 95% CI: 0.096–0.306; p < 0.001). The reliance on intrinsic motivation and communal norms could prove to be more sustainable than dependence on external incentives for fostering sanitation practices. Additionally, it is important to highlight that individuals who did not consider the construction or maintenance of latrines to be costly were more likely to be ODF (OR = 1.934; p = 0.003), albeit this was not significant after adjustment (AOR = 1.526; p = 0.169). Perceived costs may initially influence the uptake of latrines, but other factors like norms and enforcement may take precedence in determining long-term behaviour. Finally, respondents from communities where the majority felt embarrassed about not having a latrine were more likely to be ODF (AOR = 1.412; p = 0.046), suggesting that collective social pressure and shared values regarding sanitation can serve as a significant motivator in achieving and maintaining ODF status, as illustrated in Table 4.

**Table 4.** Association between social norms and open defecation free status Suna West Sub-County, Kenya.

| Social Norms | ODF<br>n (%) | OR (95% CI) | p value | AOR (95% CI) | p value |
| --- | --- | --- | --- | --- | --- |
| **Subjection to laws and penalties** |  |  |  |  |  |
| Yes | 60 (24.3) | Ref |  | Ref |  |
| No | 70 (51.1) | 3.256 (2.090-5.074) | <0.001 | 4.145 (2.203-7.802) | <0.001 |
| **Follow-ups and support** |  |  |  |  |  |
| Yes | 115 (33.3) | Ref |  | Ref |  |
| No | 15 (50.0) | 1.985 (1.089-3.620) | 0.025 | 2.111 (0.827-5.387) | 0.118 |
| **Expectation of rewards/Incentives** |  |  |  |  |  |
| Yes | 60 (22.6) | Ref |  | Ref |  |
| No | 70 (58.8) | 0.239 (0.150-0.380) | <0.001 | 0.172 (0.096-0.306) | <0.001 |
| **Construction/maintenance expensive** |  |  |  |  |  |
| Yes | 70 (28.5) | Ref |  | Ref |  |
| No | 50 (49.5) | 1.934 (1.251-2.991) | 0.003 | 1.526 (0.835-2.788) | 0.169 |
| **Embarrassing to see people defecate in open** |  |  |  |  |  |
| Yes | 91 (32.0) | Ref |  | Ref |  |
| No | 39 (41.9) | 1.587 (0.982-2.567) | 0.06 | 0.489 (0.194-1.232) | 0.129 |
| **Majority ashamed for not having latrine** |  |  |  |  |  |
| Yes | 86 (29.8) | Ref |  | Ref |  |
| No | 41 (45.6) | 1.927 (1.188-3.126) | 0.008 | 1.412 (1.014-5.739) | 0.046 |
| **Okay to defecate in rivers/bushes/dams** |  |  |  |  |  |
| Yes | 84 (28.2) | Ref |  | Ref |  |
| No | 46 (56.1) | 0.254 (0.151-0.427) | <0.001 | 0.623 (0.329-1.179) | 0.146 |

## DISCUSSION

### Open defecation status

This study found 66.1% reversion one-year post-ODF in previously certified households. Similarly, an earlier study had shown that 70% of villages reverted back to open defecation, three years after certification among seven sub-counties featured in the study (12). However, low reversion rates have been observed (8%) in Ethiopia and Ghana after one year of CLTS implementation and (14.5%) in Indonesia after two years of ODF certification (4; 10). In these studies, latrine presence - latrine presence and usage – was used as the measure for sustainability, the possible reasons for recording lower reversion rate. Nevertheless, reversion has been found to be common in villages within sub-Saharan Africa where it has been associated with several factors (17, 9) Moreover, sustainability of ODF achievements has been previously found to be a major challenge in Kenyan communities.

In a study by Tyndale-Boscoe et al. (2013) in Uganda, Kenya, Ethiopia and Sierra Leone two years after CLTS, a 13% reversion was reported when latrine presence was used to measure sustainability. However, the reversion rate would have drastically increased to 92% had the study used the 5 indicators used during the initial verification process which included functional latrine, means of keeping flies away (water seal or squat hole cover), absence of faecal matter, presence of hand washing facility with soap/ash and evidence of latrine use in the re-verification (18). This, is a much higher reversion rate like the 66.1% observed in this particular study when all the indicators are used to measure sustainability.

While governments and most organisations have been very successful in getting households to build and retain latrines, less success has been achieved in improving sanitation behaviour change which is the major aim of CLTS (18). Overall, the findings of this study suggest that there is need to harmonise or standardize indicators of ODF status. Furthermore, although the current protocol is very clear on the non-negotiable indicators, there is need to re-look at their role in defining ODF status. Doing so will help in defining concepts upfront, in developing any kind of monitoring tool of post-ODF status.

### Association between sanitation hygiene practices and open defecation status

We found significant association between sanitation hygiene practices and ODF status. Further, demonstrated that households that complied with the sanitation hygiene practices were more likely to be ODF. This resonates with studies done in Indonesia in which participants from better performing villages on ODF outcomes reported that messages around sanitation promotion and good hygiene had been constantly promoted through mosques and churches. Further, local groups carried out monitoring after CLTS implementation in an effort to promote hand washing with soap, treating of drinking water, proper food handling, solid and liquid waste management by households(10).

That households that poured ash in the pit latrines were found to be ODF is not surprising because pouring of ash in the pit latrines manages smell from latrines and therefore encourages consistent latrine use by all members of the household. This finding supports findings of previous study in which it was reported that smelly and unimproved latrines turned people back to open defecation in Ethiopia(19). Further, latrine usage by women was tampered with negatively as a result of perceptions around latrine cleanliness and smell inside (10). While this particular study found that 21% of households presented no evidence of the use of a hand-washing facility and 46% households did not wash their hands with soap and water always after using a latrine, in a study done in four African countries, there was an overall reversal rate of 17% for signs of use of a handwashing facility and 75% for consistent handwashing with soap and water. Reversion for consistent hand washing with soap and water in Homa Bay and Kilifi stood at 83% and 67%(18). Thus, this study recorded much lower reversal rate on consistent hand washing with soap as compared to previous studies.

Training on hand washing with soap, water treatment, preparing food in a hygienic way and proper storage and solid wastes disposal are standard parts of sanitation program (20). According to Lilje, in a study done in Chad, the individual perception to treating water was rated high. Respondents thought positively about the issues of water treatment and did not perceive it to be taking much effort, time or cost (21). This mirrors the findings of this present study, households that treated water were high. Water treatment commodities were available in public health offices and distributed by CHVs at household level during dry seasons, other commodities were offered at health facilities to mothers attending clinics and further, there were chlorine dispensers strategically situated in communal water points.

### Association between social norms and open defecation status

The study found association between a given number of normative and empirical expectations and ODF status a reflection of the existence of social norms within Suna West Sub County. Indeed, the existence of social norms is predicated on their functions, which in turn are driven by ecological expectation (11). This notion is supported by the results of this study. However, a potential limitation of the current study is whether such norms have broader functions needed to support programmes such as CLTS beyond their active phase.

Results of this study found that household that responded that the health of the family motivates them to be ODF were less likely to be ODF. This is in spite of previous studies like that done by UNICEF that reported that the most prominent motivator towards ODF status was concern for the health of the family(17). Households believed that stopping open defecation resulted in reduction in diarrheal diseases thus motivating them to stop open defecation (14). In another study, Moran, (2017) reports that health, even though may not have been a driver for the initial defecation behaviour change, people do continue to make effort to maintain and use latrine because of their health and that of the family(22).

Further, the study found no association between care for the family, latrine accessibility and ODF status a reflection of no prudential personal normative belief and no association was found in privacy/security offered by latrine and even peer pressure and ODF status. In a previous study, provision for privacy for superstructure, pride and the convenience of using of latrines were found to be important drivers for women in respect to building latrines in Indonesia (10). This particular study found an correlation between those that were embarrassed about not having a latrine and ODF status. This is in tandem with previous studies that reported that shame/ disgust motivated households into behaviour change(14).

This study found that individuals who did not consider the construction or maintenance of latrines to be costly were more likely to be ODF, however, this was not significant after adjustment to indicate that perceived costs may initially influence the uptake of latrines, but other factors may take precedence in determining long-term behaviour change. This is consistent with previously studies that reported high cost of building, maintenance and repair of latrines were among the reasons for reversion to non-ODF status (19). In their study, Bongartz et al. (2016) suggested that though CLTS was a zero-subsidy strategy, there was need for incorporation of sanitation marketing to CLTS to help those who can afford make informed choice even though this could pose challenge of interfering with behaviour change process (23).

Those that said rewards and incentives motivate them to be ODF were found to be less likely to be ODF. This resonates with the findings of a study done in East Java in which households that received some form of subsidy did not become ODF, it was discovered that subsidy was divisive since it was never enough for all households and thus hampered collective action, also, incentives has been found to have the capacity to corrupt intrinsic motivation (19; 11). In the study done on sustainability by UNICEF (2014), some of the enablers of sustainability were natural leaders working together and post-ODF follow up by CHVs (14)

Novotný et al. (2017), in their study, concluded that social norms were important instrumentally as sanitation outcomes depended on the level to which social influences were able to shape the perceptions of benefits or risks on sanitation-related awareness in positive ways(8). Similarly, Odagiri et al. (2017) found that in addition to economic levels and lack of reliable access to water, weaker social norms were significantly associated with the reversion to open defecation practices (10). When looked at singly, latrine usage and open defecation were sustained meaning the social sanctions played out well. However, in the other areas of hand washing with soap, provision of privacy and use of squat hole cover, there was significant reversion registered meaning the social sanctions weren’t applied across all the non-negotiable indicators. Suna West sub county had no deep-rooted social norms neither did the CLTS process inculcate new norms to bring about the overall change in sanitation hygiene practices desired and to sustain it after the pressure of certification was off.

## CONCLUSION

There was partial reversion to non-ODF status in households one year after certification. This was mainly attributed to 3 major indicators: provision of hand washing facility, squat hole cover and privacy. Moreover, there was sustained ODF status in households with good sanitation hygiene practices. It was, however, evident that social norms were not embedded on the CLTS process, thus failing to create social norms around sanitation and hygiene practices to enhance community collective action towards ODF status sustainability. Therefore, it is important to enhance good hygiene sanitation practices, while instilling social norms to inspire collective and sustainable action needed in programmes such as CLTS.

## Data Availability

The data has been made available along with the manuscript.

## ACKNOWLEDGEMENT

We acknowledge the people of Suna West Sub-County without whom this research would not have been done.

## AUTHORS CONTRIBUTION

N.R.A. conceived the presented idea. N.R.A. developed and performed the study under the supervision of C.O.A. and P.O.O. All authors discussed the results and contributed to the final manuscript.

